# Sports Related Concussion Impacts Speech Rate and Muscle Function

**DOI:** 10.1101/2020.06.14.20130443

**Authors:** Russell E. Banks, Deryk S. Beal, Eric J. Hunter

## Abstract

**Objective:** To examine speech rate and muscle function in athletes with and without sports related concussion (SRC).

**Methods:** We recruited 30 athletes aged 19-22 years-old who had sustained a SRC within the past 2 years and 30 pair-wise matched controls with no history of SRC from the student community at Michigan State University. Speech rate and muscle function were evaluated during diadochokinetic (DDK) tasks. Speech rate was measured via average time per syllable, average unvoiced time per syllable, and expert perceptual judgement. Speech muscle function was measured via surface electromyography over the obicularis oris, masseter, and segmental triangle. Group differences were assessed using MANOVA, bootstrapping and predictive ROC analyses.

**Results:** Athletes with SRC had slower speech rates during DDK tasks than controls as evidenced by longer average time per syllable (F(1, 52) = 11.072, p =.002, [95% CI : .01 to .04]), longer average unvoiced time per syllable (F(1, 52) = 16.031, p < .000, [95% CI : .01 to .029] and expert judgement of slowed rate (F(1, 22) = 9.782, p = .005, [95% CI : .163 to .807]). Rate measures were predictive of concussion history. Further, athletes with SRC required more speech muscle activation than controls to complete the DDK tasks (F(1, 3) = 17.12, p =.000, [95% CI: .003 to .006]).

**Conclusion:** We found clear evidence of slowed speech and increased muscle activation during the completion of DDK tasks in athletes with SRC histories relative to controls. Speech rate and muscle assessment should be incorporated into clinical evaluation of concussion.

## INTRODUCTION

Speech production is a complex neuromotor process dependent on millisecond timing and coordination of a large number of cortical and subcortical structures and muscle groups^1–4^. The temporal features of speech motor control are particularly sensitive to subtle changes in neural function and have been leveraged as early indicators of neurological impairment associated with Parkinson’s disease^5^, Huntington’s disease^6,7^, Amyotrophic Lateral Sclerosis^8,9^, Chronic Traumatic Encephalopathy (CTE)^10,11^ and traumatic brain injury (TBI)^12,13^. Speech timing changes may also be an important early indicator of sports-related concussions (SRC), a subset of TBI.

To the untrained listener, changes in speech timing and coordination are often perceived as “slurred speech.” Slurred speech is widely recognized by contact sport referees as an on-field sign that an athlete has sustained a SRC^14^. The most frequently cited sideline assessments and guidelines list slurred speech as a physical sign of acute SRC and encourage that speech be monitored at follow-up^15–18^. Surprisingly, no operational definition of slurred speech as it pertains to SRC has been established and no protocol exists to assess speech motor control in SRC. Establishing a meaningful and efficient speech evaluation to inform current comprehensive SRC assessments could improve diagnostics and prognostics for affected athletes, as underscored by the call for speech analyses in SRC by the American Medical Society for Sports Medicine’s (AMSSM)^17^.

Diadochokinetic (DDK) speech tasks allow for the easy and efficient evaluation of speech timing attributes^19–21^. Rate of production and pause time for DDK speech tasks can be measured objectively and subjectively using short recordings to inform measurements of an athlete’s speech characteristics post SRC. Speech rate during DDK tasks has shown promise for distinguishing SRC from controls^21^. Preliminary findings from a small mixed sample of patients with SRC and non-SRC mild TBI found slower DDK rates in patients compared to controls^13^. The present study builds on this preliminary evidence to comprehensively quantify DDK speech rate across objective and subjective measures and to examine, for the first time, any articulatory muscle differences using surface electromyography (sEMG), in athletes with SRC compared to controls. We aimed to determine a clinically relevant protocol for the evaluation of speech production and speech muscle physiology in the context of SRC.

## METHODS

### Enrollment

Under protocol approval from the Michigan State University Human Research Protection Program, all 60 participants were recruited from a student research participation pool from the University’s College of Communication Arts and Sciences. Potential participants were selected for further screening based on survey responses to determine their eligibility for inclusion in this study. The 30 participants with SRC (aged 19-22 years) were current or former athletes, with English as their first language, and a negative self-reported history of dyslexia and neurological disorders. The participants with SRC sustained their most recent concussion within the last 2 years, as motor function may be affected long-term in this population^22^. Participant \ must have been diagnosed by an MD or athletic trainer, had not been hospitalized for their injury, and had not lost consciousness for more than 20 minutes at the time of injury. One SRC participant’s data was excluded from analysis after the participant later reported the presence of an exclusionary co-morbid neurological disorder. The 30 control (CON) participants were recruited as pairwise matches for the SRC group for height, weight, education, age and gender. See Appendix I for demographic data of our sample.

### Patient and Public Involvement

Participants were not involved in this research to comment on the study design, interpret the results, contribute to the writing or editing of the manuscript.

### Procedures and Analyses

#### Speech Tasks

Participants performed a series of speech and oromotor tasks after being fitted with a microphone and sEMG sensors over the obicularis oris, masseter, and segmental triangle (see *Recording Equipment* below). The speech DDK tasks were three Alternating Motion Rate (AMR) tasks on the individual syllables *puh, tuh*, and *kuh;* and three Sequential Motion Rate (SMR) tasks on *puh-tuh-kuh*; and the words *pattycake* and *buttercup* (see Table 1). Tasks were demonstrated for clarity. Participants were instructed to produce DDKs at their maximum rate. Participants practiced the DDK tasks before recording.

**Table 1.** List of speech tasks and the timing, perceptual and sEMG analyses.

| Tasks | Objective Analysis | Subjective Analysis |
| --- | --- | --- |
| **DDK REPETITIONS:**<br><u>AMR syllables</u> - *puh*, *tuh*, *kuh* syllables (repeated 10 times each)<br><u>SMR syllables non-word</u> - *puh-tuh-kuh* (repeated 10 times)<br><u>SMR real words</u> - *pattycake* & *buttercup* (repeated 10 times each) | **ACOUSTIC TIMING:**<br><u>Average time per syllable</u> - *TPS*: [total time to produce 10 repetitions in seconds / 10 repetitions]<br><u>Average unvoiced time per syllable</u> - *UTPS*: [(total time to produce 10 repetitions in seconds – total voicing time in 10 repetitions) / 10 repetitions]<br>**sEMG:**<br><u>Average sEMG (RMS)</u> - over 10 repetitions | <u>Articulatory Precision</u> – “Are they producing the target syllable?”<br><u>Articulatory Rate</u> – “Compared to a ‘normal’ modulus, is speech faster or slower?”<br><u>Rhythmic Consistency</u> – “Is the length of each syllable and the time between syllables uniform?” |

#### Recording Equipment

The speech audio signals were recorded using an omnidirectional head-mounted microphone (M80, Glottal Enterprise, Syracuse, NY, USA) connected to a digital audio recorder (Roland R-05, 44,100 Hz, 24-bit, wav file). The microphone was placed 3-4 cm from the side of the mouth (outside the primary airstream). A wireless micro sEMG system (Trigno; www.delsys.com/products/wireless-emg) was used to detect speech motor muscle activation and electrical activity. A PowerLab 8/35 (ADInstruments, New South Wales, Australia) interfaced with the sEMG system and the digital audio recorder to simultaneously sample all signals which were recorded on a laptop computer running LabChart 10.1 software. The sEMG sensors (see Figure 1) were attached to the skin on the obicularis oris (OO), masseter (MAS), and submental triangle (ST; where tongue muscle activation can be measured) with manufacturer-supplied hypoallergenic adhesives. Sensor placement was guided by standard research practices for recording both normal and disordered speech^23–25^. Prior to sEMG placement, the location was prepared per standard practices (e.g. cleaned, lightly abraded)^23,25^.

**Figure 1.**
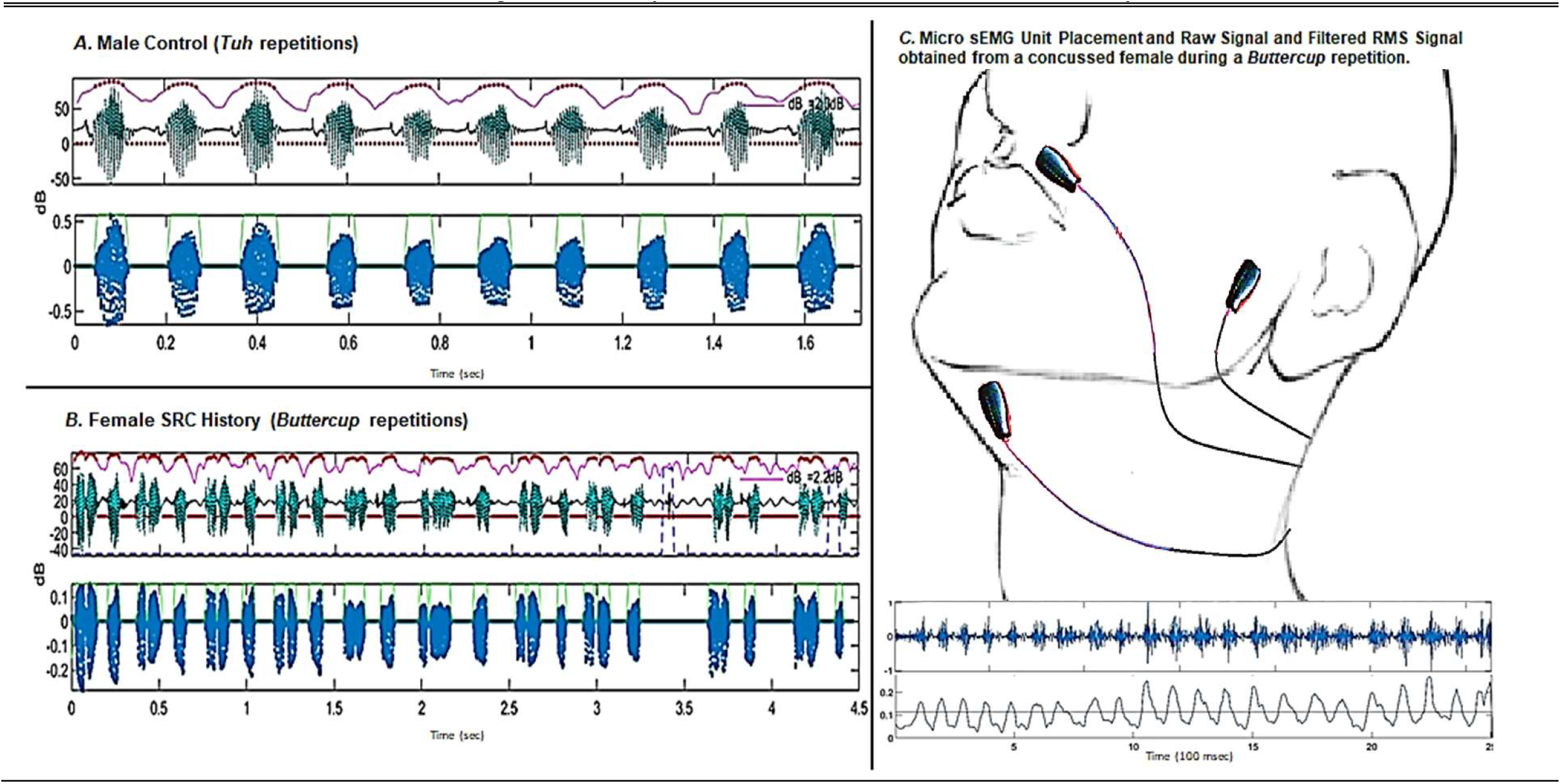
Sample acoustic waveform and timing results obtained from control male (A.) and SRC history female (B. & C.) using automated timing and sEMG analysis. The green wave forms in figures A and B are the unfiltered acoustic signal while the blue waveforms are the filtered signal. The space between green lines on the blue waveforms indicates the estimated voiced time. Figure 1C. also depicts the placement of sEMG units over the obicularis oris, massater, and submental triangle and the position of the head-mounted microphone.

### Objective Speech Measures: Timing

We created three schedules for task completion, each with its own randomized order, and then randomly assigned a schedule to each participant with SRC. The control participants were given the same schedule as the participant with SRC with whom they were matched. All participants completed three practice attempts of each task to establish familiarity with the protocol and ensure correct performance. Segmentation was performed in Audacity audio manipulation software (https://www.audacityteam.org/) and analyzed using an automated MatLab script following general guidelines described by Salvatore et al and other recent publications^13,26,27^. Briefly, this analysis applied a dB and frequency filter between 5-75 dB that speech is most likely to be present during this study (Figure 1). The automated analyses were all manually reviewed for accuracy by the first author of this experiment. Three sets of ten correct repetitions were counted during each AMR and SMR task using audio recordings and, when necessary, spectral and wave form settings.

#### Estimated Average Time Per Syllable (TPS)

Final segmentations included the 2^nd^ -11^th^ correctly produced target^13^. These ten productions were used in the statistical analyses described below. Timing measures for each set of ten repetitions were then averaged and used in statistical analysis. These segmented recordings became the basis for all timing, acoustic and perceptual analyses. To calculate the average time per production, the total length of each file containing the ten repetitions of each DDK task was divided by the total number of syllables. Only correctly articulated multisyllabic DDK productions were included in our time by count analysis.

#### Average Unvoiced Time Per Syllable (UTPS)

The length of unvoiced time was calculated as the time in which voicing was not detected during DDK productions divided by the number of syllables (10 for the AMR’s and 30 for the SMR’s). Speech timing analysis was automated using a Matlab script described in a recent publication.

### Objective Speech Measures: Surface Electromyography Measures

#### Root Mean Squared (RMS-sEMG)

We used RMS-sEMG to quantify the amount of electrical activity in the motor units of the muscles under study. There is a positive linear relationship between EMG output and imposed load of facial muscles^28,29^. This measure was applied to muscles in the jaw, lip and base of tongue in the current study to determine potential differences in electrical activity of these muscles between groups during the performance of DDK tasks. Direct current signal offset was filtered so that signal means were zero. A bandpass filter was applied between 20 and 450Hz and windowed at 500ms in order to remove movement and equipment artifact and noise (Figure 1)^25^.

### Perceptual Ratings

#### Articulatory Precision and Rhythmic Consistency

We used defined measures of articulatory precision and rhythmic consistency to rate the accuracy and precision of concussed speakers^30^. Measures were explained to the expert speech-language pathologist (SLP) raters.

#### Rate

Previous research found changes in basic parameters of the voices of individuals following a concussion, primarily in their fundamental frequencies and variability^31,32^. However, no study to date has described a formal subjective evaluation of rate in this population. It was anticipated that trained SLP listeners would note changes in rate in those with SRC histories compared to speech of those without SRC histories. SLP judges were provided a “normal” modulus to compare to each recording in terms of rate, articulatory precision, and rhythmic consistency (see Table 1).

## RESULTS

### Objective Speech Rate Analysis

A MANOVA using average times per syllable (TPS) and unvoiced times per syllable (UTPS) of all DDK tasks as independent variables was completed. Mauchly’s Test of Sphericity revealed the sphericity assumption was violated for each of the timing variables used in this model (p<.001 for each variable), therefore a Greenhouse-Geisser correction was applied. The model showed a between-subjects main effect of SRC on measures of TPS (F(1, 52) = 11.072, p =.002, *np2* =.18 [95% CI : .01 to .04]) and UTPS (F(1, 52) = 16.031, p < .000, *np2* = .24 [95% CI : .01 to .029].

To determine which specific DDK tasks were significantly different between groups, bootstrapping (5,000 reshuffles) of control and SRC data was performed and results are reported in Table 2. Results showed slower average TPS for those with a history of SRC compared to controls during all DDK productions. Additionally, longer average UTPS was noted during all DDK tasks except *kuh (p= 0*.*051) and pattycake (p= 0*.*135)* productions (Table 2). Figure 2 illustrates mean differences, effect sizes and confidence intervals in average TPS and UTPS using Gardner-Altman plots^33^. Figures 2*A* and 2*B* illustrate differences in times per syllable and unvoiced times per syllable, respectively.

**Table 2.**
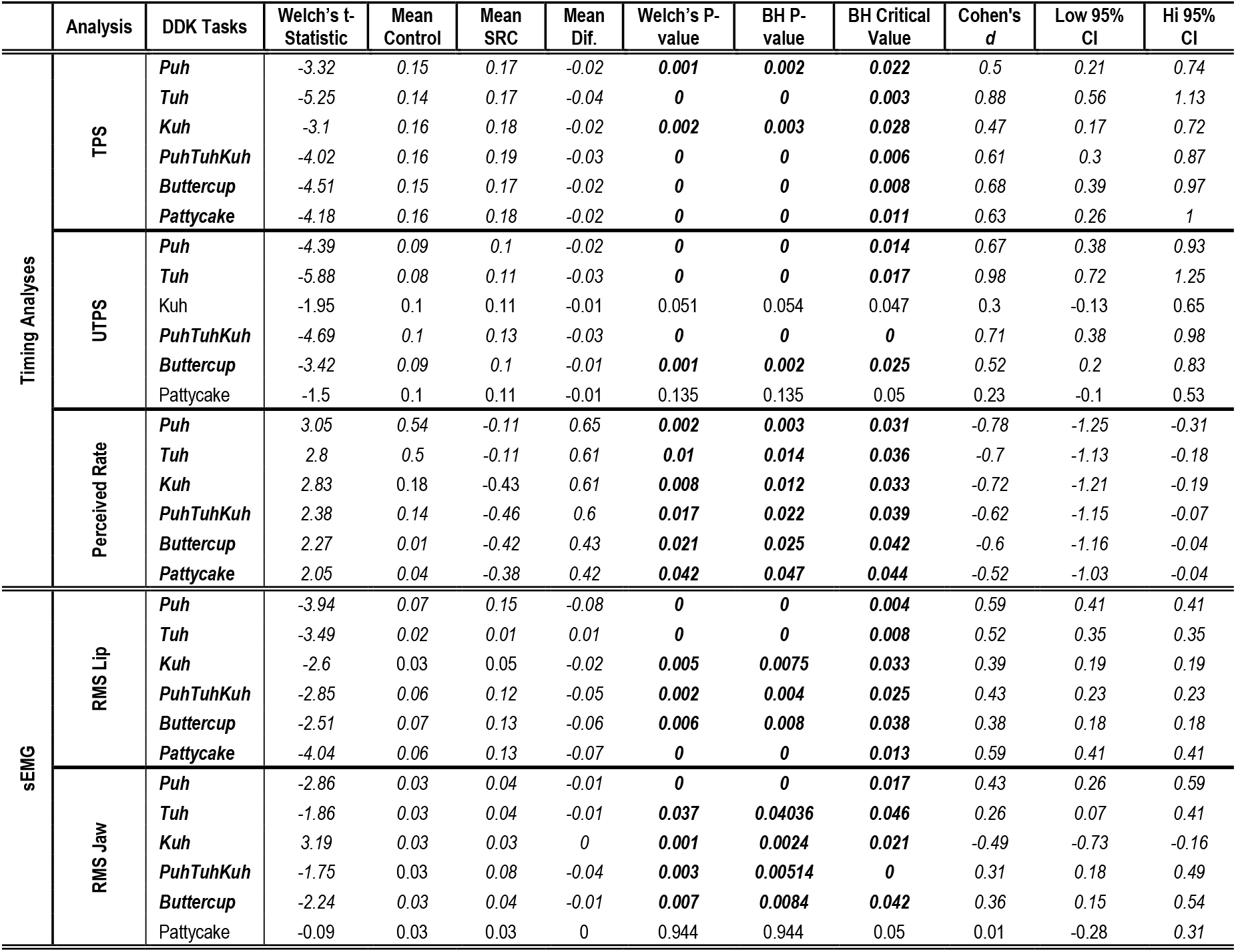
Bootstrapping results with Welsch’s t-statistic provided for timing analyses and sEMG data obtained during DDK tasks demonstrated clear timing differences between SRC and control groups. Means are reported in syllables per second. Negative mean differences indicate that syllable times for those with a history of concussion were longer than those with no concussion history. TPS = time per syllable; UTPS = unvoiced time per syllable. A Benjamini-Hochberg (BH) correction was applied where corrected P-values below the BH critical values .047 represents significant results.

|  | Analysis | DDK Tasks | Welch's t-Statistic | Mean Control | Mean SRC | Mean Dif. | Welch's P-value | BH P-value | BH Critical Value | Cohen's d | Low 95% CI | Hi 95% CI |
| --- | --- | --- | --- | --- | --- | --- | --- | --- | --- | --- | --- | --- |
| Timing Analyses | TPS | *Puh* | -3.32 | 0.15 | 0.17 | -0.02 | **0.001** | **0.002** | **0.022** | 0.5 | 0.21 | 0.74 |
|  |  | *Tuh* | -5.25 | 0.14 | 0.17 | -0.04 | **0** | **0** | **0.003** | 0.88 | 0.56 | 1.13 |
|  |  | *Kuh* | -3.1 | 0.16 | 0.18 | -0.02 | **0.002** | **0.003** | **0.028** | 0.47 | 0.17 | 0.72 |
|  |  | *PuhTuhKuh* | -4.02 | 0.16 | 0.19 | -0.03 | **0** | **0** | **0.006** | 0.61 | 0.3 | 0.87 |
|  |  | *Buttercup* | -4.51 | 0.15 | 0.17 | -0.02 | **0** | **0** | **0.008** | 0.68 | 0.39 | 0.97 |
|  |  | *Pattycake* | -4.18 | 0.16 | 0.18 | -0.02 | **0** | **0** | **0.011** | 0.63 | 0.26 | 1 |
|  | UTPS | *Puh* | -4.39 | 0.09 | 0.1 | -0.02 | **0** | **0** | **0.014** | 0.67 | 0.38 | 0.93 |
|  |  | *Tuh* | -5.88 | 0.08 | 0.11 | -0.03 | **0** | **0** | **0.017** | 0.98 | 0.72 | 1.25 |
|  |  | *Kuh* | -1.95 | 0.1 | 0.11 | -0.01 | 0.051 | 0.054 | 0.047 | 0.3 | -0.13 | 0.65 |
|  |  | *PuhTuhKuh* | -4.69 | 0.1 | 0.13 | -0.03 | **0** | **0** | **0** | 0.71 | 0.38 | 0.98 |
|  |  | *Buttercup* | -3.42 | 0.09 | 0.1 | -0.01 | **0.001** | **0.002** | **0.025** | 0.52 | 0.2 | 0.83 |
|  |  | *Pattycake* | -1.5 | 0.1 | 0.11 | -0.01 | 0.135 | 0.135 | 0.05 | 0.23 | -0.1 | 0.53 |
|  | Perceived Rate | *Puh* | 3.05 | 0.54 | -0.11 | 0.65 | **0.002** | **0.003** | **0.031** | -0.78 | -1.25 | -0.31 |
|  |  | *Tuh* | 2.8 | 0.5 | -0.11 | 0.61 | **0.01** | **0.014** | **0.036** | -0.7 | -1.13 | -0.18 |
|  |  | *Kuh* | 2.83 | 0.18 | -0.43 | 0.61 | **0.008** | **0.012** | **0.033** | -0.72 | -1.21 | -0.19 |
|  |  | *PuhTuhKuh* | 2.38 | 0.14 | -0.46 | 0.6 | **0.017** | **0.022** | **0.039** | -0.62 | -1.15 | -0.07 |
|  |  | *Buttercup* | 2.27 | 0.01 | -0.42 | 0.43 | **0.021** | **0.025** | **0.042** | -0.6 | -1.16 | -0.04 |
|  |  | *Pattycake* | 2.05 | 0.04 | -0.38 | 0.42 | **0.042** | **0.047** | **0.044** | -0.52 | -1.03 | -0.04 |
| sEMG | RMS Lip | *Puh* | -3.94 | 0.07 | 0.15 | -0.08 | **0** | **0** | **0.004** | 0.59 | 0.41 | 0.41 |
|  |  | *Tuh* | -3.49 | 0.02 | 0.01 | 0.01 | **0** | **0** | **0.008** | 0.52 | 0.35 | 0.35 |
|  |  | *Kuh* | -2.6 | 0.03 | 0.05 | -0.02 | **0.005** | **0.0075** | **0.033** | 0.39 | 0.19 | 0.19 |
|  |  | *PuhTuhKuh* | -2.85 | 0.06 | 0.12 | -0.05 | **0.002** | **0.004** | **0.025** | 0.43 | 0.23 | 0.23 |
|  |  | *Buttercup* | -2.51 | 0.07 | 0.13 | -0.06 | **0.006** | **0.008** | **0.038** | 0.38 | 0.18 | 0.18 |
|  |  | *Pattycake* | -4.04 | 0.06 | 0.13 | -0.07 | **0** | **0** | **0.013** | 0.59 | 0.41 | 0.41 |
|  | RMS Jaw | *Puh* | -2.86 | 0.03 | 0.04 | -0.01 | **0** | **0** | **0.017** | 0.43 | 0.26 | 0.59 |
|  |  | *Tuh* | -1.86 | 0.03 | 0.04 | -0.01 | **0.037** | **0.04036** | **0.046** | 0.26 | 0.07 | 0.41 |
|  |  | *Kuh* | 3.19 | 0.03 | 0.03 | 0 | **0.001** | **0.0024** | **0.021** | -0.49 | -0.73 | -0.16 |
|  |  | *PuhTuhKuh* | -1.75 | 0.03 | 0.08 | -0.04 | **0.003** | **0.00514** | **0** | 0.31 | 0.18 | 0.49 |
|  |  | *Buttercup* | -2.24 | 0.03 | 0.04 | -0.01 | **0.007** | **0.0084** | **0.042** | 0.36 | 0.15 | 0.54 |
|  |  | *Pattycake* | -0.09 | 0.03 | 0.03 | 0 | 0.944 | 0.944 | 0.05 | 0.01 | -0.28 | 0.31 |

**Figure 2.**
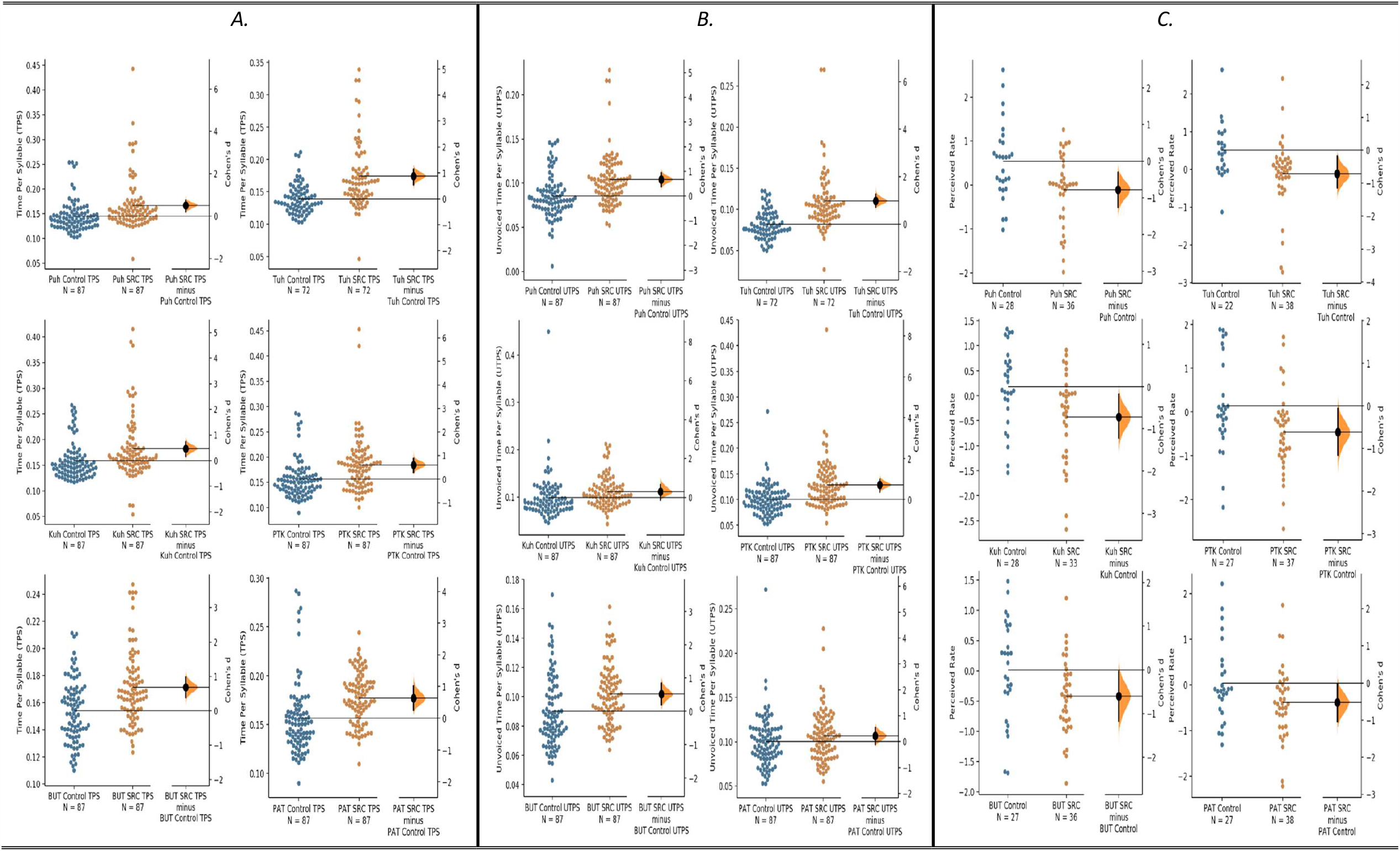
The average TPS (*A*), UTPS (*B*), and perceived rate (*C*) for all DDK comparisons are shown in the above Gardner-Altman estimation plots. Control data is plotted in blue and SRC history plotted in brown; each Cohen’s d is plotted on a floating axis on the right as a bootstrap sampling distribution. Cohen’s d is depicted as a black dot; bootstrap 95% confidence intervals are indicated by the ends of the black vertical error bars.

### Perceptual Analysis

A MANOVA was performed using raters’ judgments of articulatory precision, articulatory rate, and rhythmic consistency for all six DDK tasks as independent variables. No violations to the assumptions were noted in this model. A between-subjects main effect of SRC history on trained SLP judgements of speech rate (F(1, 22) = 9.782, p = .005, *np2* =.308 [95% CI : .163 to .807]). The model did not detect a between-subjects main effect of SRC history on raters judgments of articulatory precision (F(1, 22) = .326, p =.574, *np2* =.015 [95% CI : -.603 to .343]) or rhythmic consistency (F(1, 22) = .141, p = .710, *np2* = .006 [95% CI :CI : -.32 to .416]). To determine which specific DDK tasks were perceived as significantly different between groups, bootstrapping (5,000 reshuffles) of control and SRC data was performed and results indicated that all DDK tasks were perceived to be significantly slower than controls (Table 2. & Figure 2C.).

### SEMG ANALYSIS

#### Root Mean Squared (RMS)

A MANOVA was performed analyzing the average measured output of EMG units attached to the face during the production of all DDK tasks. The model showed a between-subjects main effect of SRC history on the average measures EMG output across muscles of the face (F(1, 3) = 17.12, p =.000, *np2* =.118 [95% CI: .003 to .006]. To further investigate which DDK tasks produced significant differences, bootstrapping analyses of lip, jaw, and tongue muscle activation were performed comparing SRC and controls.

#### Lip

The model showed a between-subjects main effect of SRC history on the average measures RMS-EMG output of the obicularis oris. Individuals with a history of SRC demonstrated significantly greater lip muscle activation compared to participants with no SRC history during all DDK tasks with the exception of *tuh* productions, where controls had significantly more lip muscle activation (Table 2 & Figure 3 *A*).

**Figure 3.**
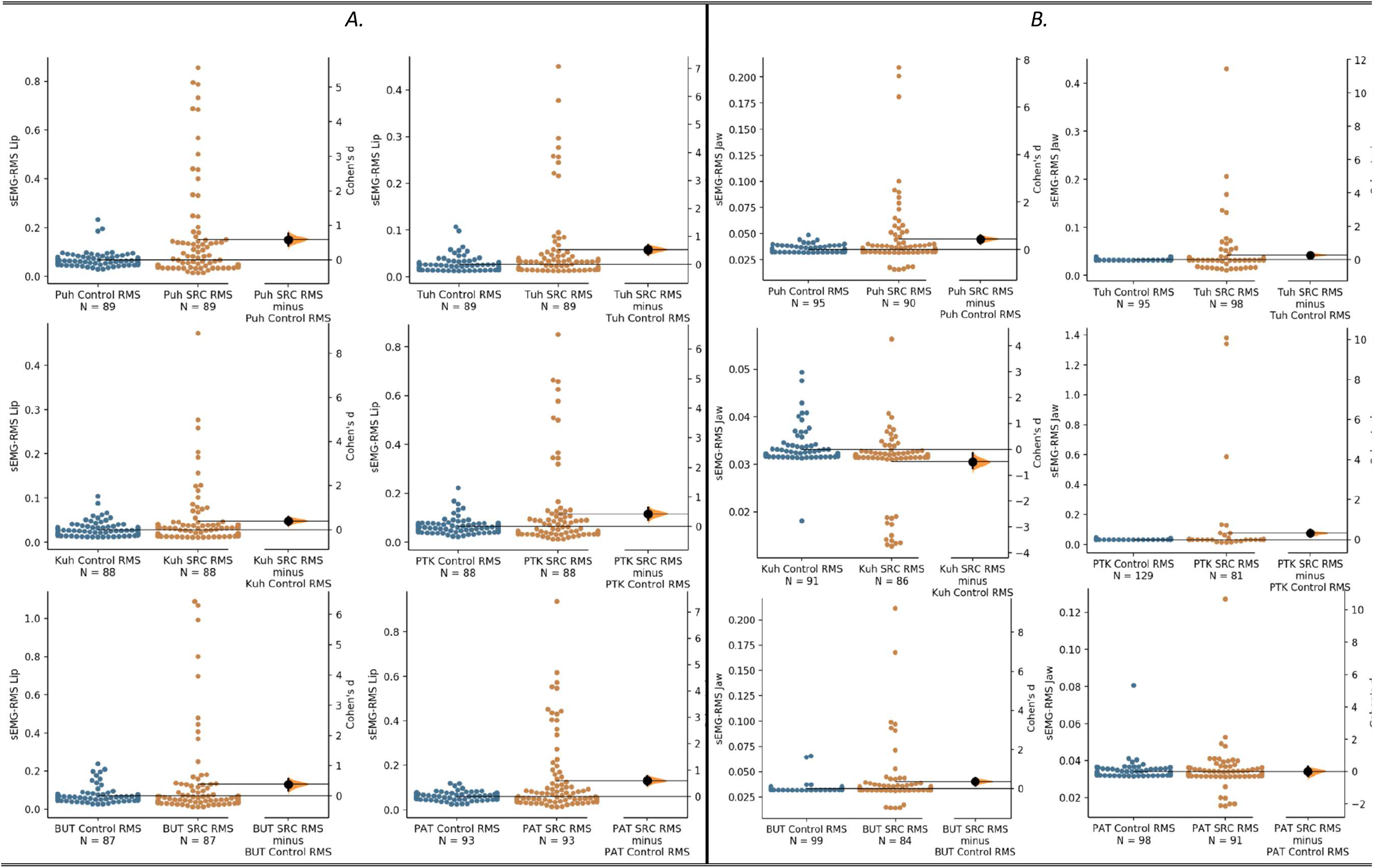
The average sEMG-RMS of Lip (*A*) and Jaw (*B*) muscle activation during all DDK comparisons are shown in the below Gardner-Altman estimation plots. Control data is plotted in blue and SRC history plotted in brown; each Cohen’s d is plotted on a floating axis on the right as a bootstrap sampling distribution. Cohen’s d is depicted as a black dot; bootstrapped 95% confidence intervals are indicated by the ends of the black vertical error bars.

#### Jaw

The model showed a between-group difference on the average measures of RMS-EMG where those with a history of SRC demonstrated significantly greater muscle activation in the masseter muscles of participants with a SRC history compared to controls during all DDK productions with the exception of *pattycake* (p=.0944) productions only (Table 2. & Figure 3 *B*).

#### Tongue

The model was unable to detect differences between groups on the average measures of RMS-EMG where those with a history of concussion demonstrated greater muscle activation in the base of tongue compared to participants with no SRC history.

### AUROC Analysis

An area under the receiver operating curve (AUROC) analysis was carried out to understand the predictive diagnostic ability of each of the above measures (see Figure 4). The predictive value of our TPS measure to identify patients with concussion indicates *Tuh* productions possess the highest discriminatory ability (moderate at Area Under the Curve (AUC)=0.77). Assuming values greater 0.136 syllable/sec indicate a positive concussion history, sensitivity was high (0.86) but specificity was low (0.54). For our UTPS measure, *Puh* had the highest discriminatory value (AUC= 0.83). With a cut-off of 0.0844 seconds/syllable, sensitivity was high (0.83) and specificity moderate (0.7). SLP’s Perception of rate during *Buttercup* provided the highest discriminatory value (AUC= 0.75). Applying a cut-off of approximately seven perceived units slower than the provided modulus, sensitivity was high (0.84) but specificity was low (0.62). AUROC values obtained for sEMG data were not able to reliably distinguish between groups.

**Figure 4.**
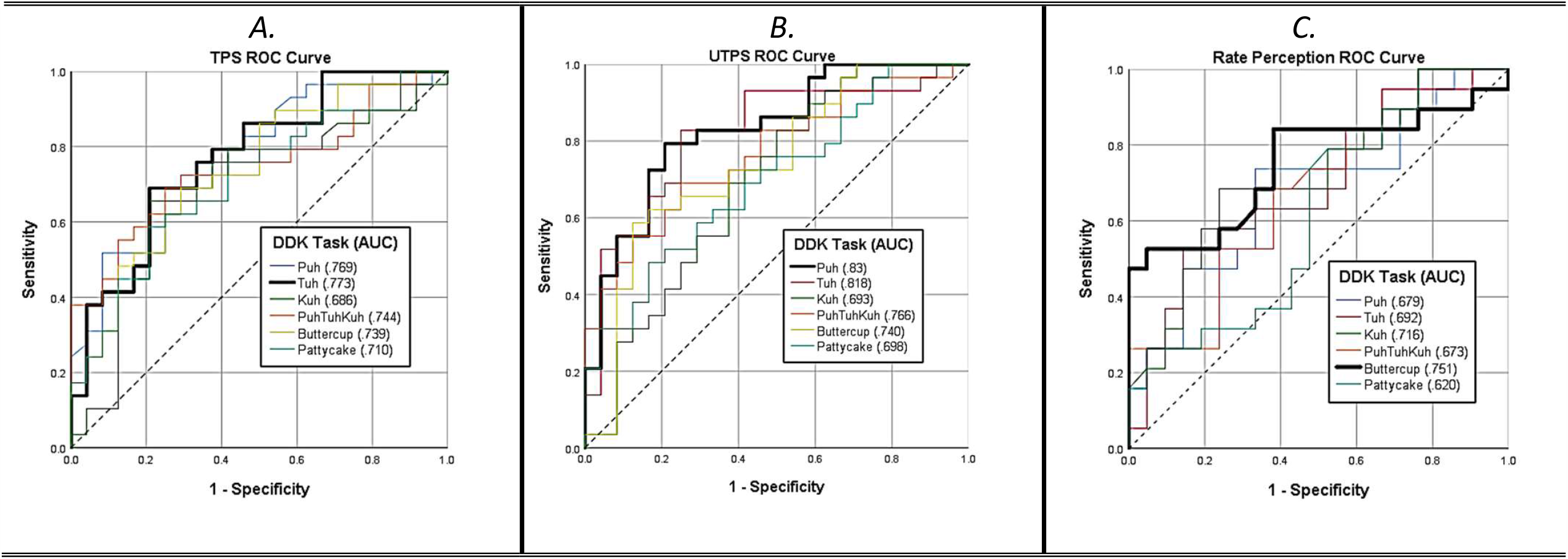
AUROC for TPS (*A*), UTPS (*B*), and perceived rate (*C*) for all DDK tasks with associated Area Under the Curve (AUC) values. Dark black lines indicate those tasks for each measure which were most readily able to distinguish between groups of positive or negative concussion histories. Hashed lines are provided as the reference (.50) indicating no predictive value.

## DISCUSSION

The speech rate of athletes with SRC histories was slower during DDK tasks compared to controls. Athletes with SRC histories produced all DDK’s slower and with longer unvoiced time (pauses) per syllable compared to controls. Corroborating objective findings, eight trained SLP’s evaluated the speech of athletes with SRC histories to be slower than controls across DDK tasks. Further, both objective and subjective measures of rate had good discriminatory power for athletes with SRC history. Finally, athletes with SRC demonstrated mostly increased lip and jaw muscle activation during the production of DDK tasks as compared to controls.

### Speech Rate

With the potential for SRC to cause widespread neurometabolic dysregulation, the observed motor speech deficits may be related to cortical motor dysfunction akin to the early motor speech deficits indicative of ALS, Parkinson’s Disease and CTE^5,8– 10,34,35^. For example, speech rate decreases and pause increases, may be potentially sensitive in identifying those in the early stages ALS^36^. Further, speech articulatory rate and pause measures have been used to identify individuals in early stages of

Parkinson’s disease^34,37^. Perhaps most intriguing are the implications that our results have for our understanding of CTE. CTE is characterized postmortem as degradation and toxic accumulation of pathogens in the cortical and subcortical structures responsible for cognitive and motor function. This degradation is likely multifactorial including repeated concussive and sub concussive head injury^10,11^. To date, the studies that classify CTE speech patterns have not included younger, former, nonprofessional athletes with SRC histories, but instead, described older, former, professional athletes with suspected (self-reported) CTE who suffer from a characteristic disease and symptom profile. Concerningly, we identified that the speech rates of former nonprofessional athletes with supposedly resolved concussion symptomology, that is, no diagnosis of progressive disease related to repeated head injury, was clearly slowed when compared to healthy matched controls for simple speech DDK tasks.

Normative DDK rate values have been established for healthy non-brain injured English speakers (ages 15-65) for the token *puhtuhkuh* at 6.2 syllables/second. Itch and Ben-David suggest that values of 5.4 syllables/second and slower may be potentially pathological and require further evaluation^21^. Healthy controls in our study produced 6.3 *puhtuhkuh* syllables/second (±.035) compared to 5.4 syllables/second (±.048) for those with SRC histories, a rate considered pathological. Further, we found that DDK rate measures had fair to excellent discriminatory power with good sensitivity to reliably predict concussion history status. Given the minimal time and low cost of adding a DDK rate task to all concussion assessment time points, the high sensitivity and moderate specificity is acceptable. Further investigation will aim to increase the specificity of our measures.

SLP perceptual evaluations of speech were consistent with the findings of the effect of SRC on objective timing analyses, where participants with SRC histories were rated as clearly slower during DDK repetitions than controls. Together with the objective timing evidence presented in this study, it is reasonable to deduce that participants with SRC histories did, in fact, produce clearly slower DDK repetitions. Additionally, SLP’s were reliably able to detect differences between groups with and without concussion histories, particularly during *Buttercup* repetitions. Thus, with proper training, speech professionals may be sensitive to speech differences in this population.

### sEMG Analysis

Novel application of speech articulator muscle engagement analysis using sEMG provides additional evidence of the negative effects of SRC history on speech musculature function. Participants with SRC histories demonstrated increased muscle activation compared to controls on speech tasks routinely requiring the engagement and coordination of lip and jaw musculature. RMS-EMG results of articulatory speech muscle output described in this work are consistent with findings of increased cortical motor excitability due to increased neurotransmitter release and NMDA (excitatory neurotransmitter) receptor activation in individuals with SRC^38,39^. This increase may be partially responsible for the decreased speech rate during DDKs tasks, resulting in increased rigidity and decreased articulatory function.

## CONCLUSIONS

Long-lasting deficits in objective speech timing, physiological articulatory muscle power, and subjective evaluations of rate exist between individuals with SRC histories compared to those without. Given that participants were no longer experiencing SRC related symptoms, we anticipate that analyses presented in this work may be used to identify individuals with acute (active symptomology) and non-acute SRC, persistent concussive syndrome, and the potential severity of SRC injuries. Our findings have clinical implications as measures established in this work may have diagnostic and prognostic utility, be used in a mobile fashion, and be sensitive to changes over time which can inform return to play recommendations. Future work will apply the motor speech analysis methods outlined in this study to predict and track SRC recovery, degenerative disease, and potential risk factors associated with long-term outcomes.

## Data Availability

All data applicable to the research in this manuscript are available upon request for further analysis, in the event that analysis does not interfere with current data analysis projects.

## Acknowledgments

We would like to thank all the members of the Michigan State University Voice Biomechanics and Acoustics Laboratory and especially Dr. Lady Cantor-Cutiva for her statistical consultation.

## Ethics Approval

This Speech Analysis of Former Concussed Athletes study was approved by the institutional review board of human research projects at Michigan State University and carried out in accordance with Michigan States University’s standards of ethics.

